# Cognitively Healthy Centenarians are genetically protected against Alzheimer’s disease specifically in immune and endo-lysosomal systems

**DOI:** 10.1101/2023.05.16.23290049

**Authors:** Niccolo’ Tesi, Sven van der Lee, Marc Hulsman, Natasja M. van Schoor, Martijn Huisman, Yolande Pijnenburg, Wiesje M. van der Flier, Marcel Reinders, Henne Holstege

## Abstract

**BACKGROUND:** Alzheimer’s Disease (AD) prevalence increases with age, yet a small fraction of the population reaches ages beyond 100 years without cognitive decline. We aimed to uncover the genetic factors associated with such resilience against AD.

**METHODS:** Genome-Wide-Association-Studies (GWAS) identified 86 single-nucleotide-polymorphisms (SNPs) associated with AD-risk. We studied each SNP in 2,281 AD-cases, 3,165 middle-aged population controls, and 346 cognitively healthy centenarians, and we combined SNPs into Polygenic Risk Scores (PRS) for each individual. Finally, we investigated the functional properties of the SNPs enriched/depleted in centenarians using *snpXplorer*.

**RESULTS:** Centenarians were depleted with risk-increasing AD-SNPs and enriched with protective AD-SNPs. The PRS was more than 5-fold lower in centenarians compared to AD cases (*p*=7.69×10^−71^) and almost 2-fold lower compared to middle-aged population controls (*p*=5.83×10^−17^). The strongest protection was found in *ANKH, GRN, TMEM106B, SORT1, EPDR1, PLCG2, RIN3, CD2AP*, and *APOE* associated alleles. As expected, the genetic protection was diluted in the offspring of the centenarians.

**DISCUSSION:** Becoming a cognitively healthy centenarian is associated with a complex genetic protection against AD, which concentrates on an advantageous functioning of the endo-lysosomal and immune systems, and their effect on amyloid-clearance.

## Introduction

The average human life expectancy continues to grow and by 2050 there will be 3.2 million centenarians in the world. [1] At old ages, a major contributor to poor health is cognitive decline and dementia, of which Alzheimer’s Disease (AD) is the most common type. [2, 3] However, AD is not an inevitable consequence of aging, as testified by a small proportion of the population that reaches at least 100 years while maintaining a high level of cognitive and physical functions. [4, 5] This raises the question of whether these centenarians have exceptional features that protect or delay the onset of dementia, and whether such mechanisms may be genetically encoded.

AD is a progressive disorder characterized by loss of cognitive functions, ultimately leading to loss of independence and death, for which an effective treatment is lacking. [3, 6] The greatest risk factor for AD is age: the disease is rare at 60 years, and the incidence of AD reaches ∼40% per year at 100 years of age. [7] Next to aging, heritability plays an important role which changes dramatically with age. While the heritability of AD with age at onset <65 years is estimated to be 90%-100%, mostly due to autosomal dominant or strong risk-increasing genetic variants, [8] it decreases to 60-80% for ages at AD onset of ∼75 years (from twin studies), based on a unique mix of rare and common risk factors, and further declines with later ages at AD onset. [9] Approximately 30% of the genetic risk of AD is attributable to the e4 allele of the *APOE* gene. Large collaborative Genome-Wide Association Studies (GWAS) have collectively identified 84 additional Single Nucleotide Polymorphisms (SNP) that are associated with a slight modification of the risk of AD. [10, 11]

Intriguingly, the reverse is also true, since ∼60% of the chance to survive to 100 years in good cognitive health depends on inheriting favorable genetic factors, [12] comprising a relative depletion of risk-increasing variants and an enrichment of advantageous genetic variants that associate with a prolonged (brain) health. [13–15] In fact, we previously found that using cognitively healthy centenarians as controls in a case-control study of AD, rather than controls that are age-matched with the AD cases, led to an average twofold increase in effect size of the association of AD-SNPs. [16] Consequently, cognitively healthy centenarians had a significantly lower Polygenic Risk Score (PRS), compared to AD cases and healthy normal controls, based on 29 previously identified AD-association risk SNPs.

In this study, we further explore whether our findings when investigating the 86 SNPs that were recently identified to affect the risk of AD. [10] We studied the effect of individual SNPs on developing AD as well as their combined effect (PRS). Furthermore, we identified the risk-increasing and protective SNPs that were respectively most depleted or enriched in a cohort of cognitively high-performing centenarians, which allowed us to highlight the biological mechanisms most strongly involved with resilience against Alzheimer’s disease. Last, we investigated to what extent the favorable genetic constellation was observable in the genomes of the (adult) children of the centenarians.

## Methods

### Cohort description

We included 6,747 individuals in this study. Of these, 2,542 were AD cases, either clinically diagnosed with probable AD from the Amsterdam Dementia Cohort (N=2,060) [17] or pathologically confirmed from the Netherlands Brain Bank (N=482). [18] Next to AD cases, we used 3,643 subjects as healthy controls, age-matched with the AD cases. Finally, we used 360 cognitively healthy centenarians and 202 children of the centenarians, from the 100-plus Study cohort. [5] Additional information regarding the cohorts is available in *Supplementary Methods: Populations*. The Medical Ethics Committee of the Amsterdam UMC approved all studies. All participants and/or their legal representatives provided written informed consent for participation in clinical and genetic studies.

### Genotyping and imputation of 86 selected SNPs

We included 86 SNPs (plus SNP rs12459419 near *CD33*) [19, 20] that were significantly associated with AD in the latest GWAS by *Bellenguez* et al., (*Table S1*). [10] After quality control and genotype imputation of the genetic data (see *Supplementary Methods: Genotyping and Imputation*), all individuals passed quality control. Before analysis, we excluded individuals of non-European ancestry (based on 1000Genomes clustering) [21] and individuals with a family relation (identity-by-descent ≥ 0.2), [22] leaving 2,281 AD cases, 3,165 healthy controls, 346 centenarians, and 193 centenarian-children for the analyses.

### Single variant analyses

As reference effect size for each SNP, we used the effect sizes resulting from the comparison of 39,106 clinically-diagnosed AD cases and 401,577 age-matched controls used in the discovery phase by *Bellenguez* et al., (*Table S1*). [10] We excluded the proxy phenotypes which *Bellenguez* et al., included in their multi-stage meta-analysis, as these are based on paternal and maternal disease status rather than clinical diagnosis, which typically leads to a dilution of the SNP effect sizes. For each AD-associated SNP, we calculated the *change* in effect size relative to the reference effect size when comparing (i) AD cases *vs*. centenarians, (ii) AD cases *vs*. age-matched healthy controls, and (iii) healthy controls *vs*. centenarians (see *Supplementary Methods: Change in effect size*).

### Polygenic Risk Score

We combined all 86 SNPs into a Polygenic Risk Score (PRS), resembling an individuals’ net genetic risk of AD. As weights for the PRS, we conventionally used the effect sizes of the meta-analysis including both clinically-diagnosed AD cases and by-proxy phenotypes, reflecting the final results of *Bellenguez* et al., (*Table S1*). Given the large effect size associated with the two *APOE* SNPs (rs429358 and rs7412), we calculated PRS including and excluding these two SNPs. We assessed the association between PRS and AD risk by comparing the scaled PRS (μ=0, σ=1) between AD cases, healthy controls, and centenarians in a pairwise manner. Then, we validated the associations by comparing the centenarians’ children with age-matched healthy controls: to do so, we used all available children (possibly related) as well as a set of unrelated children. For the associations, we used logistic regression models adjusting for population stratification (PC1-5). The resulting effect sizes (log of odds ratio) can be interpreted as the odds ratio difference per one standard deviation increase in the PRS, with the corresponding 95% confidence intervals.

### The contribution of a centenarian

In genetic studies, the power to detect a significant SNP-association is influenced by the effect-size and the number of cases and controls in the comparison. The larger the effect-size, the lower number of cases and controls needed for an association to reach statistical significance. We have previously shown that using centenarians as controls in a case-control study of AD led to a twofold enrichment of the effect-size of AD-associated SNPs. Here, we formalize this power increase by estimating the number of normal controls each centenarian is worth. To do so, for each SNP identified by *Bellenguez* et al., we calculated the number of normal (age-matched) controls and cognitively healthy centenarians necessary to obtain 80% power to find a SNP association at p-value = 0.05. We assumed (i) 8,000 AD cases, (ii) the minor allele frequency as reported in the reference GWAS (*Table S1*), and (iii) the observed effect size from our comparisons (AD cases vs. age-matched controls, and AD cases vs. cognitively healthy centenarians). Because the direction of effect *must* be consistent with the direction reported in *Bellenguez* et al., we excluded SNPs for which we observed an opposite direction of effect in both AD cases *vs*. age-matched controls and AD cases *vs*. cognitively healthy centenarians. Then, for each SNP, we compared 8,000 AD cases with 200 controls, and recursively increased the number of controls by 200 until a power of at least 80% was found or the number of controls was twice the number of AD cases (*i*.*e*., 16,000). When a SNP-association reached at least 80% power, we regarded it as *converging*. The ratio between the number of normal healthy controls and cognitively healthy centenarians, for each SNP, indicates the increase in statistical power of a single centenarian relative to age-matched controls. We simulated the analysis using several thresholds for the number of AD cases to use (2,500, 5,000, 8,000, and 10,000) and found that after 8,000 no additional SNPs converged.

### In silico functional analysis

We investigated the biological pathways associated with the SNPs with the largest effect-size differences between centenarians and age-matched controls. We selected SNPs for which, based on our power analysis, the number of centenarians was at least *half* of the number of age-matched controls to achieve the same power. For the functional analysis, we used the functional annotation section of snpXplorer web-server with default settings. [23] This tool performs (i) variant-to-gene mapping using integrating variant consequences (coding, intronic, intergenic) and quantitative-trait-loci (eQTLs and sQTLs), followed by (ii) gene-set enrichment analysis and (iii) clustering of the enriched terms. [23] The clusters of enriched terms were compared to clusters obtained from a previous study including all AD-associated SNPs based on the same method. [23]

### Implementation

Quality control of the genotype data, population stratification analysis, and relatedness analyses were performed with *PLINK* (v1.90 and v2.0). Association analyses, downstream analyses, and plots were performed with R (v4.2). To estimate the number of normal controls each centenarian is worth we adapted the likelihood ratio test framework implemented in the R package *genpwr*. [24] The scripts are publicly available at https://github.com/TesiNicco/Centenarians_AD.

## Results

### Quality control of genetic data and SNPs

The mean age at study inclusion of the 2,281 AD cases was 67.96 ± 9.84 (55% females), the mean age of the 3,165 healthy controls was 62.57 ± 8.66 (48% females), the mean age of the 346 centenarians was 101.05 ± 2.51 (71% females), and the mean age of the 193 centenarians’ children was 70.31 ± 8.64 (53% females) (*Table 1*). The median quality of the imputed SNPs was r2=0.95 and ranged 0.45-0.99 (*Table S2*); all imputed SNPs were included in the analyses.

**Table 1.**
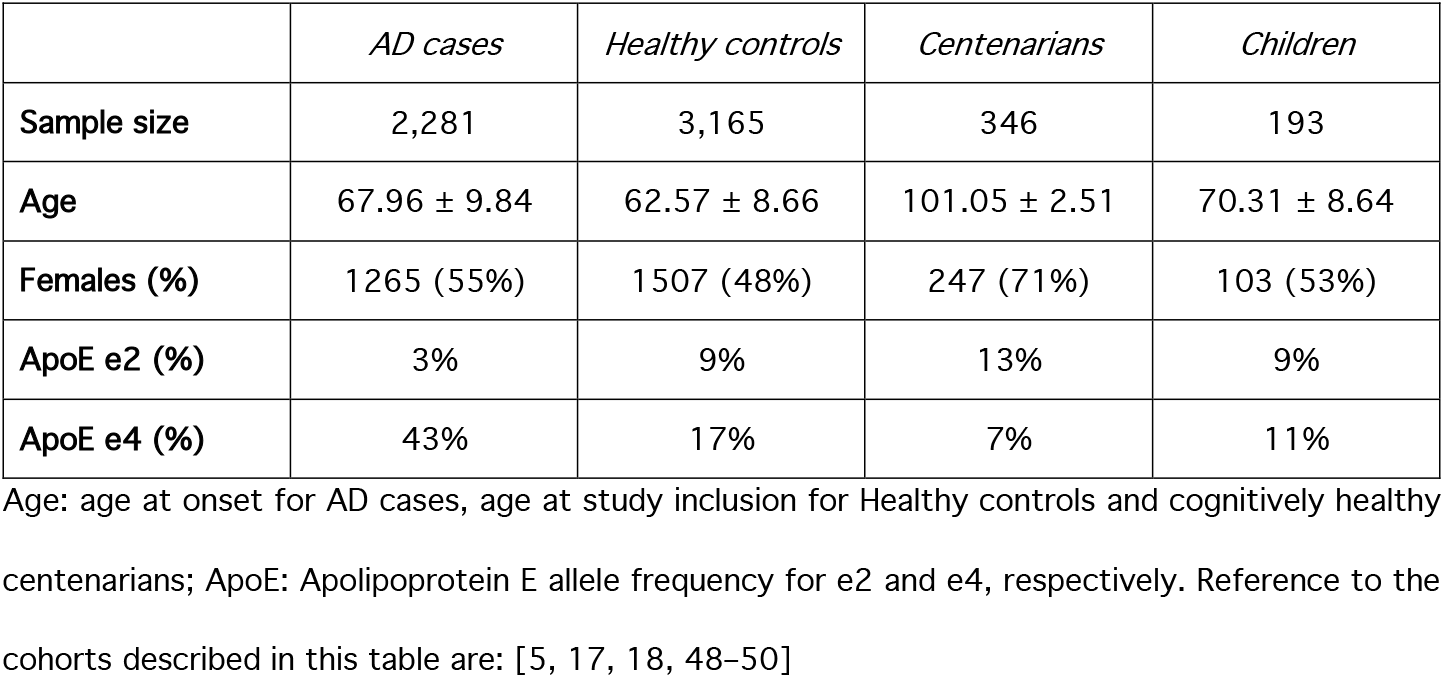
Population characteristics

|  | *AD cases* | *Healthy controls* | *Centenarians* | *Children* |
| --- | --- | --- | --- | --- |
| **Sample size** | 2,281 | 3,165 | 346 | 193 |
| **Age** | 67.96 ± 9.84 | 62.57 ± 8.66 | 101.05 ± 2.51 | 70.31 ± 8.64 |
| **Females (%)** | 1265 (55%) | 1507 (48%) | 247 (71%) | 103 (53%) |
| **ApoE e2 (%)** | 3% | 9% | 13% | 9% |
| **ApoE e4 (%)** | 43% | 17% | 7% | 11% |

Age: age at onset for AD cases, age at study inclusion for Healthy controls and cognitively healthy centenarians; ApoE: Apolipoprotein E allele frequency for e2 and e4, respectively. Reference to the cohorts described in this table are: [5, 17, 18, 48–50]

### AD cases vs. Centenarians

When comparing AD cases with cognitively healthy centenarians, the effect size across all 86 tested SNPs increased by a median 1.78-fold (IQR: 0.51-2.85) relative to the published effect sizes; *Figure 1*, Figure S1, *Table S3* and *Table S4*). [10] For 59 SNPs the *change* in effect size was >1 (*p* = 3.6×10^−4^ based on a one-tailed binomial test, *Figure 1*) and ranged from 1.07 (rs785129 near *HS3ST5* gene) to 5.91 (rs112403360 in *ANKH* gene, *Table S3*). The centenarians did not include carriers of the rare rs60755019 (in *TREML2*), while the carriers-frequency in AD cases was 0.18% and 0.14% in age-matched controls (*Table S4*). For 9 SNPs (in or near the genes *EPDR1, MAF, PLCG2, RIN3, ANKH, TMEM106B, SORT1, GRN*, and *WDR12*), the effect size was increased more than 4-fold compared with previously published effect sizes (*change* > 4). The effect of 16 SNPs was not increased compared to the reference effect sizes (0 < *change* < 1, *Figure 1* and *Table S3*), and the effect of 11 SNPs was opposite compared to the reference effects (*change* < 0, *Figure 1, Figure S1* and *Table S3*). Despite the small sample size of centenarians, the association with AD reached significance after multiple test corrections for 8 out of 85 SNPs (*FDR* < 5%): *ANKH, GRN, PLCG2, RIN3, ABCA7, BIN1*, and the 2 *APOE* SNPs, *Figure 1* and *Table S3*).

**Figure 1:**
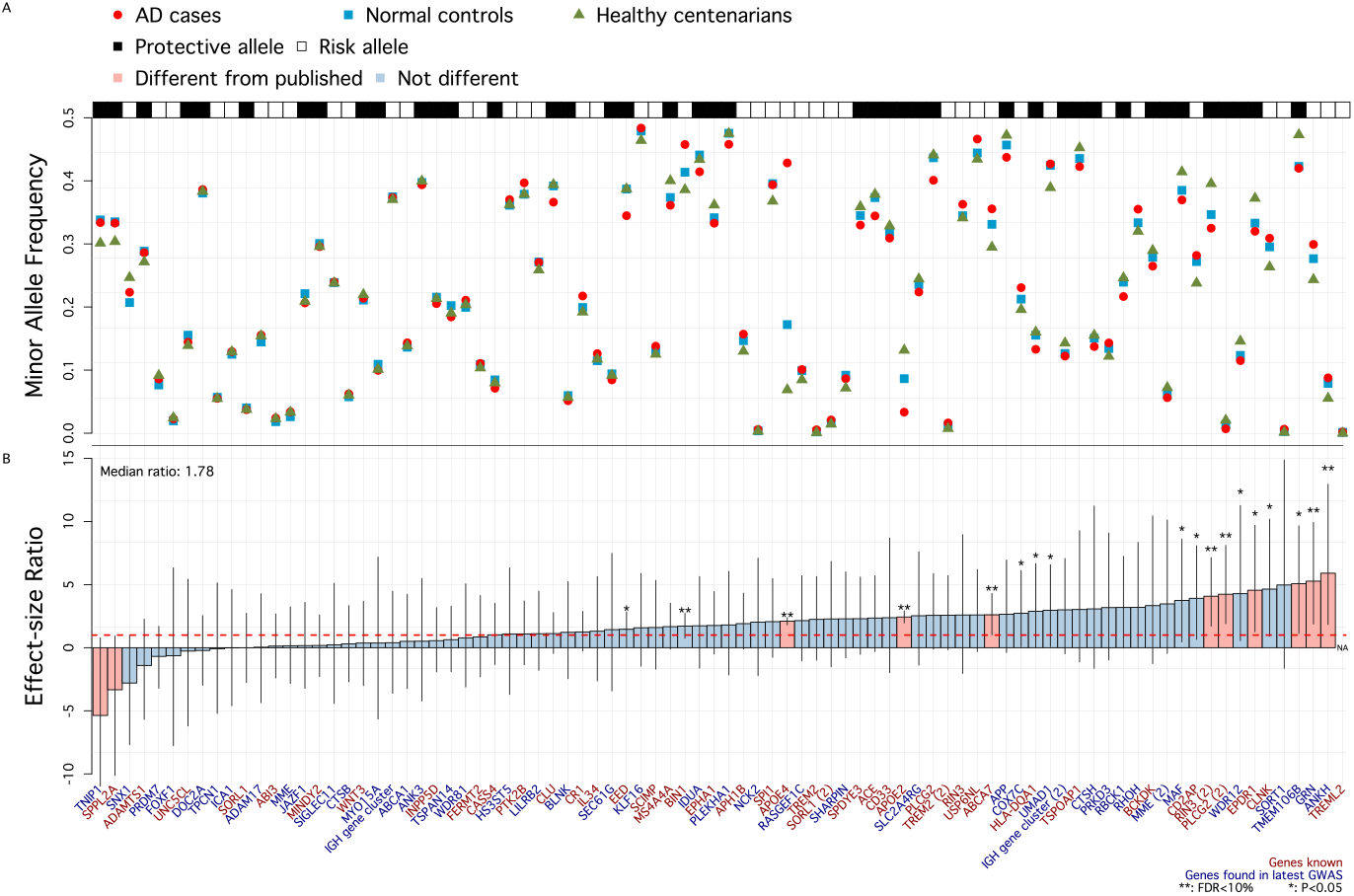
Single variant associations summary. Figure A (top) shows the raw minor allele frequency in AD patients (red circles), healthy controls (blue squares), and cognitively healthy centenarians (green triangles). Black and white annotation squares refer to whether the plotted allele (the minor allele) was associated with an increased risk of AD (Risk allele, white) or a decreased risk of AD (Protective allele, black). Figure B (bottom) shows the *change* in effect size when comparing observed effect sizes (AD cases vs. cognitively healthy centenarians) to the reference effect sizes (*Bellenguez* et al.). Blue genes refer to novel SNP-AD associations discovered by *Bellenguez et al*. for the first time, while red genes were known before *Bellenguez et al*. The dashed red line at 1 indicates the published effect size from the literature. *: *p-value* of association <0.05; **: *FDR-corrected p-value* of association < 0.05; Pink bars indicate SNPs for which observed effect size is significantly different from published effect size.

### AD cases vs. age-matched controls

The 2,281 AD patients have mainly early onset AD, such that they are likely enriched with risk-increasing genetic variants relative to the predominantly late onset AD cases included in *Bellenguez* et al. [10] Therefore, our AD dataset may explain part of the change in effect sizes observed in our AD vs centenarians analysis. To investigate the contribution of the AD cases, we compared them with 3,165 age-matched controls. We observed a 1.16-fold increased effect size relative to the published effect sizes (IQR: 0.60-1.76), which significantly lower than the 1.78-fold increased effect size in the comparison of AD cases *vs*. centenarians (*p* = 0.004 when comparing the distributions of effect size *change*, Figure *S2* and *Table S5*). The *change* in effect size was >1 for 48 SNPs and ranged from 1.01 (rs73223431 near *PTK2B* gene) to 4.47 (rs141749679 near *SORT1* gene). In total, a significant association after multiple test corrections (*FDR* < 5%) was identified for 11 SNPs, in or near *SORT1, RHOH, PLCG2, HLA-DQA1, EED, RIN3, APH1B, TREM2, BIN1*, and the 2 *APOE* SNPs, *Table S5*).

### Age-matched controls vs. Centenarians

The effect size of AD-SNPs in a comparison of age-matched controls *vs*. cognitively healthy centenarians was increased by a median 0.58-fold (IQR: -0.23-1.45) relative to the published effect sizes in *Bellenguez* et al. (*Figure S3* and *Table S6*). [10] The *change* in effect size was >2-fold for 17 SNPs, and one-to-twofold for 13 SNPs. The effect sizes of 29 SNPs were not increased compared with the reference effects, and the effect of 27 SNPs was opposite. Altogether, a significant association after multiple test corrections (*FDR* < 5%) was identified only for the 2 *APOE* SNPs, *Table S6*).

### Polygenic Risk Score

We assigned two PRSs to each subject, one including the weighted effect of all the 86 SNPs, and a second excluding the effect of the two *APOE* SNPs (*Figure 2*). Then, we compared the distribution of the PRSs between AD cases, healthy controls, cognitively healthy centenarians, and the centenarians’ children (*Figure 2* and *Table S7*). In all comparisons, the PRSs in AD cases was significantly higher (*Figure 2* and *Table S7*). AD patients *vs*. healthy age-matched controls, excluding the 2 *APOE* SNPs: *OR* = 1.54, *95% CI* = [1.45 – 1.63], *p* = 1.55×10^−47^; including *APOE* SNPs: *OR* = 2.55, *95% CI* = [2.39 – 2.72], *p* = 2.09×10^−176^). AD patients *vs*. cognitively healthy centenarians, excluding the 2 *APOE* SNPs: *OR* = 1.97, *95% CI* = [1.74 – 2.23], *p* = 2.75×10^−26^; including *APOE* SNPs, *OR* = 5.07, *95% CI* = [4.25 – 6.06], *p* = 1.54×10^−71^). We found a significantly lower PRS in centenarians compared to younger healthy controls, excluding *APOE* SNPs: *OR* = 0.77, *95% CI* = [0.69 – 0.88], *p* = 2.57×10^−5^; including *APOE* SNPs, *OR* = 0.53, *95% CI* = [0.46 – 0.62], *p* = 2.92×10-^17^. Finally, the centenarians’ children had a significantly lower PRS than the younger controls (including all children): including the *APOE* SNPs: *OR =* 0.74, *95% CI* = [0.62 – 0.88], *p* = 7.67×10^−4^, but not after excluding *APOE* SNPs (*OR =* 0.93, *95% CI* = [0.80 – 1.09], *p* = 0.38, *Table S7*).

**Figure 2:**
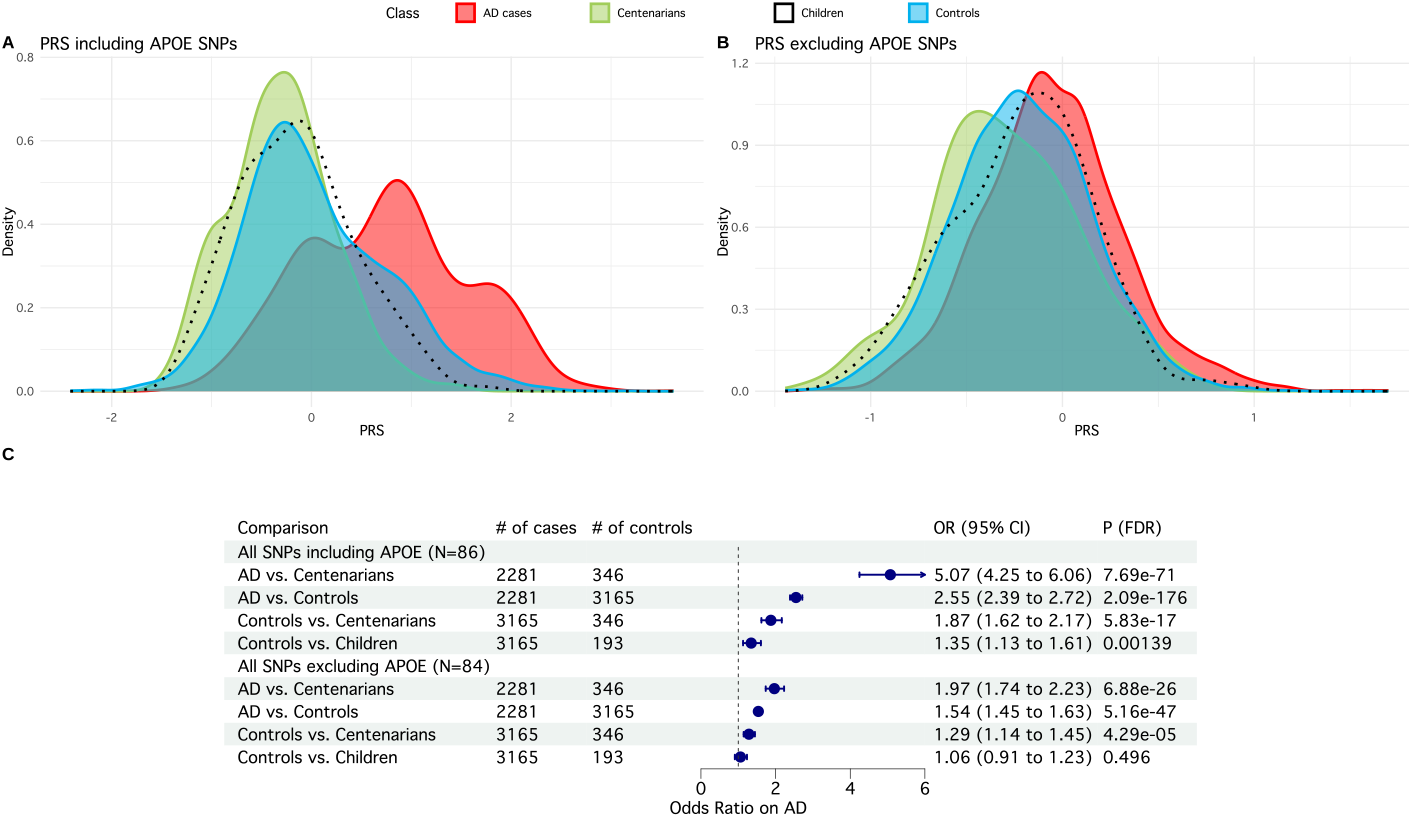
Summary of PRS including all SNPs. Figure A (top-left) shows the distribution of the PRS including the two *APOE* SNPs (86 SNPs in total) in AD cases (red), healthy controls (blue), cognitively healthy centenarians (green), and their children (transparent with black border). Figure B (top-right) shows the distribution of the PRS excluding the two *APOE* SNPs (84 SNPs in total). Figure C (bottom) shows the association statistics (Odds Ratio, 95% CI and corrected p-value) and forest plot of the PRS including and excluding *APOE* SNPs. For the comparisons, we used logistic regression models in a pairwise manner (*i*.*e*. AD cases vs. Centenarians, AD cases vs. Controls, Controls vs. Centenarians, and Controls vs. Centenarians’ children), controlling for population substructure.

### The contribution of a centenarian

To estimate the number of ‘normal’ controls and cognitively healthy centenarians required to reach 80% power to find an association at *p* = 0.05, we used a subset of 67 common SNPs for which the direction of effect in our analyses matched that of *Bellenguez* et al., (see *Methods: The contribution of a centenarian Table S3*, and *Table S8*). For 8 SNPs, a total of 16,000 controls did not guarantee the power of 80%, (*i*.*e*. no convergence) using both normal controls and centenarians, which is likely due to the small effect-sizes associated with these SNPs (*Figure 3A* and *Table S8*). For the remaining 59 SNPs, an association at *p* = 0.05 (convergence) was observed when comparing 8,000 AD cases with on average 6,183 ± 5,680 normal controls (median = 3,600, IQR = 2,300-8,800) or 3,745 ± 5,436 centenarians (median = 1,200, IQR = 600-3,300) (*Figure 3* and *Table S8*). On average, and based on 59 AD-SNPs, the power of a single cognitively-healthy-centenarian in a GWAS of AD is equivalent to that of 5.86 normal controls (median = 2.4, IQR = 1.00-6.58, *Figure 3* and *Table S8*).

**Figure 3:**
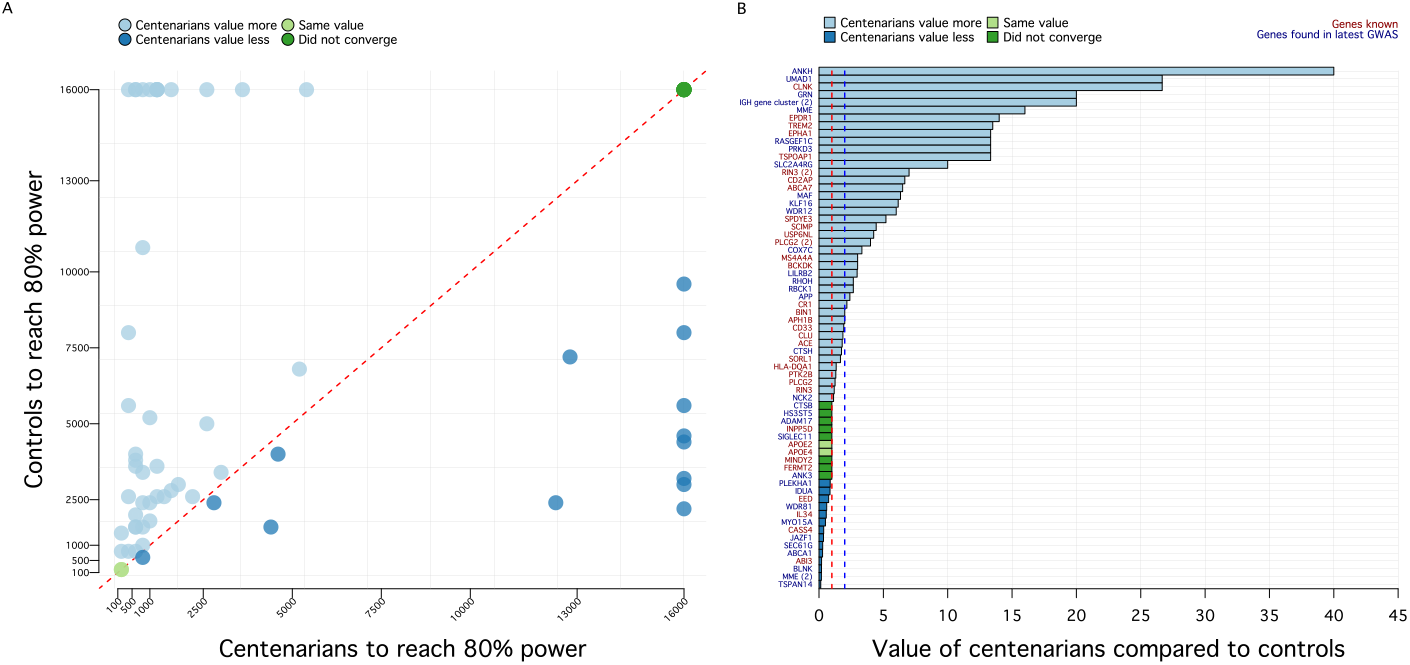
Relationship between centenarians and normal controls. Figure A (left) shows the number of individuals (normal controls on the y-axis and centenarians on x-axis) necessary to achieve 80% power for a SNP-association at p = 0.05, assuming 8,000 AD cases. We restricted this analysis to common variants (MAF>1%) with expected direction of effect in our comparisons (N=67 SNPs, see *Methods*). Note that, for this reason, some variants enriched in centenarians such as rs13237518 (*TMEM106B*) and rs13237518 (*SORT1*) could not be represented here. Each dot represents a SNP: dark green dots identify the 8 SNPs that did not converge using both normal controls and centenarians (*i*.*e*. the power did not reach 80%). Light green dots indicate the 2 *APOE* SNPs, for which N=200 individuals (normal controls and centenarians) were enough to guarantee 80% power. Light blue dots identify SNPs for which the number of centenarians (to achieve 80% power) was lower than the number of normal controls. Of these, N=13 SNPs did not converge using normal controls. Conversely, dark blue dots identify SNPs for which the number of normal controls was lower than the number of centenarians. Of these, N=8 SNPs did not converge using centenarians. Figure B (right) shows the ratio between the number of normal controls and the number of centenarians, for each SNP. Color code is the same as Figure A. SNPs larger than the blue dotted line (N=31, ratio>2) were used for functional annotation and gene-set enrichment analysis.

### Functional implications

We then functionally annotated and performed gene-set enrichment analysis using 31 SNPs for which the power of a single centenarian was >2-fold than normal controls (*Figure 3B*). Of 31 SNPs, only 2 were coding (rs143332484 in *TREM2* and rs72824905 in *PLCG2*), 23 were annotated to their likely affected gene(s) using eQTL, sQTL, and CADD information, and 6 SNPs were annotated solely based on their genomic position (*Table S9*). The resulting genes were used as input for gene-set enrichment analysis. After clustering the enriched Gene Ontology (GO) terms based on a semantic similarity measure, we found 2 clusters of pathways, pointing toward the *immune system* and *endo-lysosomal trafficking* (*Figure 4* and *Table S8*). The *immune system* cluster of pathways included *activation and regulation of immune response* (genes *CR1, MS4A6A, IGH-cluster, RIN3, KAT8, GRN, SCIMP, RBCK1, APP, RHOH, OTULIN, MAPK9, PLCG2*, and *TREM2*), *leukocyte activation and differentiation* (genes *CD55, CR1, IGH-cluster, APP, GRN, PLCG2*, and *TREM2*), *macrophage activation* (genes *GRN, APP, PLCG2*, and *TREM2*), and *neuroinflammatory response* (genes *GRN, LILRA5, PLCG2, KAT8*, and *TREM2*). The *endo-lysosomal trafficking* cluster of pathways included marked immunological aspects: *endocytosis and phagocytosis* (genes *IGH-cluster, RIN3, ABCA7, LILRB4, APP, RHOH, PLCG2*, and *TREM2*), *interleukin-6 metabolism* (genes *SCIMP, LILRA5, APP, PLCG2*, and *TREM2*), and *amyloid clearance* (genes *ABCA7, MME, APP*, and *TREM2*) (*Figure 4C, Table S8*). We compared these clusters with 5 clusters from a previous study including all AD-associated SNPs. [23] A significant overlap was found only between the *endo-lysosomal trafficking* cluster (this analysis) and (i) the *amyloid clearance* cluster (previous study, chi-square p=3.38×10^−5^), and (ii) *immune trafficking and migration* cluster (previous study, chi-square p=2.07×10^−4^). Conversely, no significant overlap was found regarding clusters of pathways pointing to *activation of immune response* (p=0.49), *organization and metabolic processes*, and *beta-amyloid and tau formation*.

**Figure 4:**
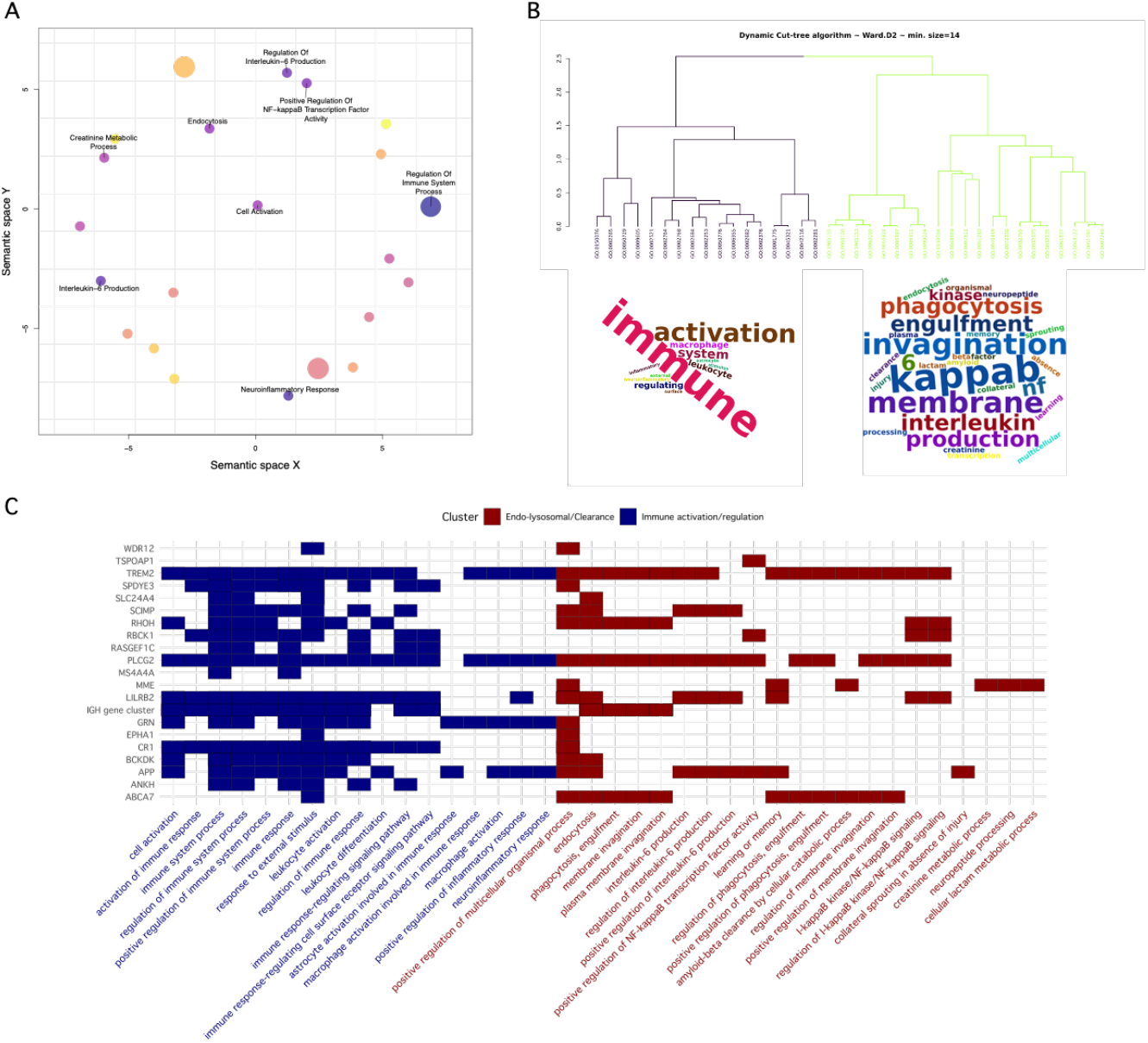
Functional annotation of SNPs with the largest effect in centenarians. The figure shows the result of the functional annotation of 31 SNPs for which the number of centenarians required to achieve 80% power was at least half of the number of normal controls required to achieve the same power. Functional annotation analysis was performed using *snpXplorer*. [23] Figure A shows the result of the gene-set enrichment analysis followed by REVIGO analysis, which clusters enriched pathways based on a semantic similarity measure. Figure B shows the dendrogram of the main enriched pathways along with their cluster (branches color code for cluster assignment) and wordclouds showing the main terms enriched in the underlying pathways. Figure C shows the mapping between significant pathways (x-axis), AD-associated SNPs (y-axis, labeled with the name of the gene as provided by *Bellenguez et al*), and the relative gene-set enrichment cluster.

## Discussion

Based on common AD-associated SNPs as identified by GWAS, self-reported cognitively healthy centenarians from the 100-plus Study are genetically protected against Alzheimer’s disease (AD) which, at least in part, explains their resilience to AD. The centenarians, and to a lesser extent their children had a significantly lower PRS for AD compared to middle-aged healthy individuals, both including and excluding the effect of the two *APOE* alleles. We observed the strongest genetic differences between centenarians and middle-aged individuals for risk alleles that are functionally associated with a specific subset of known AD-associated pathways including the endo-lysosomal system, the immune system regulation, and amyloid clearance.

Centenarians have a lower frequency of almost all risk alleles, and a higher frequency of protective alleles, as identified by GWAS, which indicates that maintaining cognitive health depends on having an advantageous function across all associated mechanisms. The effect size of each risk allele was increased by an average 1.78-fold when using centenarians as a controls rather than age-matched controls, such that one centenarian contributes the equivalent of on average 6 age-matched controls. However, some risk alleles clearly stand out, as they are more prominently depleted or enriched in centenarians than others, which suggests that some disease-associated mechanisms may be more important to maintain than others. The risk-alleles in which centenarians are most strongly depleted are the *ANKH, GRN* and *SORT1* alleles, while centenarians are most strongly enriched with the *TMEM106B, EPDR1, PLCG2* (rs72824905), and *RIN3* (rs12590654) protective alleles. For these alleles, the effect sizes were >4-fold increased when comparing AD cases with centenarians rather than age-matched controls.

Centenarians are most strongly enriched with the *ANKH* protective allele. The AD-associated risk allele in the *ANKH* gene (rs112403360) is associated with Hippocampal Sclerosis and Braak Neurofibrillary Tangles Stages. [25] Impairment of the ANKH gene leads excessive mineralization, including calcification of arteries leading to joint pain, arthritis, atherosclerosis and diabetes. [26, 27] Together, this suggests that the prolonged cognitive health in centenarians may be supported by maintained vasculature and low pathology load in brain.

Furthermore, it is intriguing that the protective alleles of the *GRN-, TMEM106B-* and the *SORT1*-associated loci are among the strongest enriched in the centenarians, as these three genes all contribute to lysosomal mechanism and endosomal trafficking. [28–30] It is notable that these loci were previously identified in context of FTLD risk. [31, 32] This might suggest that these FTLD risk alleles also influence the risk of AD, that some AD patients may have FTLD as a comorbidity, or that FTLD patients were misdiagnosed as AD patients, influencing the GWAS. [25] Regardless of rationale, the strong enrichment of these three alleles underlines the importance of a functional endolysosomal trafficking mechanism in maintained cognitive health during aging. This is further supported by a strong enrichment of the protective allele of *RIN3* (rs12590654 and rs7401792), the function of which is also associated with endo-lysosomal function and axonal trafficking. [33]

*EPDR1* (Mammalian ependymin-related protein 1) is a transmembrane protein that plays a crucial role in adhesion of neural cells. [34] Although its role in Alzheimer’s disease is currently not clear, *EPDR1* was shown to be downregulated in AD patients compared to controls, [35] and has been implicated in dopaminergic regulation of neurogenesis and neuroendocrine function in goldfish. [36] While speculative, our finding that centenarians are enriched with a protective *EPDR1* allele may confirm a role for prolonged neurogenesis in maintaining cognitive health. [37]

Protective alleles in genes modulating immune and neuroinflammatory response (*PLCG2, CR1, TREM2, OTULIN, MS4A-cluster*) were strongly enriched in centenarians, suggesting that maintaining an efficient regulation of neuro-immune response during aging is an important aspect of cognitive health. Notably, the protective coding SNP rs72824905, leading to the gain-of-function p.P522R substitution in *PLCG2*, provides proof of concept that only a limited increase in immune activation translates to a beneficial effect, since stronger gain-of-function mutations in *PLCG2* (*e*.*g*., p.S707Y and p.L848P) are associated with autoimmune disorders such as PLAID and APLAID. [13, 38, 39] *TREM2* is well-known to be involved in microglial activation and phagocytosis in the same pathway as *PLCG2*. The protective allele of the coding SNP in *TREM2* (rs75932628) leading to p.R47H, was enriched in the centenarians and was shown to increase microglial activation and expression of proinflammatory cytokines. [40] Altogether, a slightly more active immune and neuroinflammatory response in centenarians seems to better cope with the physiological accumulation of pathology over time and promote a long-term maintenance of cognitive health. [41]

The protective alleles of SNPs near *ABCA7* (rs12151021), *SORL1* (rs74685827), *APP* (rs2154481) and *APOE* (rs429358 and rs7412) were all enriched in centenarians. These genes are involved in immune-lipid signaling pathways that lead to the clearance of amyloid peptides in the brain. [42] Specifically, *ABCA7* gene is involved in beta-amyloid processing and clearance, while *SORL1* gene codes for a retromer-receptor involved in the trafficking and of amyloid precursor protein (APP), thereby preventing Aß secretion. [43, 44] Interestingly, in the brains donated by the centenarians we observed amyloid-beta deposits across many regions, however, the load of amyloid-beta neuropathology remained very limited. [45] This suggests that protective alleles in beta-amyloid clearing mechanism may associate with the resistance of amyloid-related pathology accumulation.

When we compared the AD-PRS of centenarians’ children with healthy controls, we found a significant difference only when including the high-impact *APOE* variants, of which the risk allele is are strongly depleted in centenarians. This suggests that most variants with small effects fade quickly in the offspring, and that the genetic protection that we observe in the centenarians occurs only when the majority of protectives alleles are carried. Note that common protective alleles (such as those investigated in this study) are expected to have only small effects on gene function, as impactful changes to conserved molecular mechanisms are likely to have a damaging effect. Thus, the functional effects of genetically-driven improvements of highly conserved molecular mechanisms are mostly limited, as exemplified by the limited functional effects of strongly protective alleles in *PLCG2, APP*, and *APOE*. [41, 43, 46, 47]

With the increasing number of individuals included in GWAS studies (the latest, which we used here, included ∼700,000 individuals [10]), the identification of SNPs that associate with AD with very small effect sizes becomes possible. While 84% (37/44) of the SNPs that were associated with AD for the first time in *Bellenguez* et al., had the same direction of effects in our comparisons, others had an effect in the opposite direction. While it is likely that there is insufficient power for the replication of limited associations in a small dataset like ours, we can also not rule out the effect of age: some SNPs may have a stronger effect at the early stages of the disease than at late onset, or SNPs may have pleiotropic effects.

Our study suggests that genetic comparisons of diseased individuals with those who are resilient to the disease maximizes the identified effect sizes, which allowed us to replicate the association of 8 SNPs at FDR<5% when comparing AD cases with only 360 centenarian controls, while a comparison of these AD cases with more than 10 times the number of age-matched controls allows for the replication of 11 SNPs. A disadvantage in using centenarians is that they need to be recruited one by one, which is a time consuming and expensive endeavor, and complicates obtaining a cohort of sufficient size for genetic analysis. Furthermore, cognitively healthy centenarians are not representative of the overall population: they were preselected to have escaped AD until extreme ages, such that using these individuals as controls might lead to an enrichment of SNPs that also contribute to longevity. Moreover, the centenarians in this study are all from the same (Dutch) population, such that effects in other ethnic backgrounds may be different. Lastly, we acknowledge that part of the individuals used in this study was also included in the GWAS study we used as a reference. However, these individuals represent <2% of all AD cases included in the GWAS, and <0.5% of all controls included, making their relative contribution negligible.

In summary, resilience or resistance to cognitive decline is supported by a relative depletion of deleterious genetic variants, and a concurrent enrichment of protective genetic elements. Altogether, we have shown that cognitively healthy centenarians are genetically protected against Alzheimer’s disease at the level of the single SNPs and at the PRS level. We showed that the SNPs with the largest effect in the centenarians were involved in the maintenance of effective immune and endo-lysosomal systems, possibly leading to better clearance of amyloid, and slowing cognitive decline.

## Data Availability

All data produced in the present study are available upon request to the authors.

## Acknowledgements

Niccolò Tesi is appointed at ABOARD. Sven van der Lee, Henne Holstege, Marcel Reinders and Wiesje van der Flier are recipients of ABOARD. ABOARD is a public-private partnership receiving funding from ZonMW Nationaal Dementiaprogramma (#73305095007) and Health∼Holland, Topsector Life Sciences & Health (PPP-allowance; #LSHM20106). More than 30 partners, including de Hersenstichting (Dutch Brain Foundation) participate in ABOARD (https://www.alzheimer-nederland.nl/onderzoek/projecten/aboard).

## Supplementary Methods

### Populations

#### Amsterdam Dementia Cohort (ADC)

The ADC comprises patients who visit the memory clinic of the VU University Medical Center, Amsterdam. The diagnosis of probable AD in this cohort was based on the clinical criteria formulated by the National Institute of Neurological and Communicative Disorders and Stroke – Alzheimer’s Disease and Related Disorders Association (NINCDS-ADRDA) and based on the National Institute of Aging-Alzheimer Association (NIA-AA). At baseline, all subjects underwent a standard clinical diagnostic assessment including neurological examination and standard blood tests. In addition, all subjects underwent magnetic resonance imaging, an electroencephalogram, and cerebrospinal fluid (CSF) was analyzed for most patients. Clinical diagnosis is made in consensus-based, multidisciplinary meetings. Together, this diagnostic procedure reduces the chance of misdiagnosis.

#### Healthy controls

As healthy controls, we used (*i*) a sample of 1,776 Dutch older adults from the Longitudinal Aging Study of Amsterdam, [50] (*ii*) a sample of 1,524 older adults with subjective cognitive decline who visited the memory clinic of the Alzheimer Center Amsterdam and SCIENCe project, and were labeled cognitively normal after the extensive examination, [17] (*iii*) a sample of 62 healthy controls from the Netherlands Brain Bank, [18] (*iv*) a sample of 196 individuals from the twin study, [49] and (*v*) a sample of 85 older adults from the 100-plus Study (partners of centenarian’s children). Individuals with subjective cognitive decline were followed up over time in the SCIENCe project, and only individuals who did not convert to mild cognitive impairment or dementia during follow-up were included in this study.

The 100-plus Study of Cognitively Healthy Centenarians and their family members

As cognitively healthy centenarians used a sample of N=360 individuals from the 100-plus Study cohort. [5] This study includes Dutch-speaking individuals who (*i*) can provide official evidence for being aged 100 years or older, (*ii*) self-report to be cognitively healthy, which is confirmed by a proxy, (*iii*) consent to the donation of a blood sample, (*iv*) consent to (at least) two home visits from a researcher, and (v) consent to undergo an interview and neuropsychological test battery. Next to the centenarians, we used N=202 of their children.

### Genotyping an Imputation

Genetic variants were determined by standard genotyping and imputation methods. Briefly, we genotyped all individuals using the Illumina Global Screening Array and applied established quality control methods. [51] We used high-quality genotypes in all individuals (individual call rate >99%, variant call rate >99%), individuals with sex mismatches were excluded and departure from Hardy-Weinberg equilibrium was considered significant at p<1×10^−6^. Genotypes were then lifted over to GRCh38 and prepared for imputation using provided scripts (HRC-1000G-check-bim.pl) specifying TOPMED as reference panel. [52] This script compares variant ID, strand, and allele frequencies to the TOPMED reference panel (version r2, N=194,512 haplotypes from N=97,256 individuals). Finally, all variants were submitted to the Michigan Imputation server (https://imputation.biodatacatalyst.nhlbi.nih.gov/). The server uses EAGLE (v2.4) to phase data and Minimac4 to perform genotype imputation to the reference panel (version r2).

## Change in effect size

The *change* in effect size was calculated using the same approach adopted in [16]. Briefly, the *change* refers to the ratio between the reference effect sizes and the observed effect sizes when comparing (i) AD cases with centenarians, (ii) AD cases with age-matched healthy controls, and (iii) healthy controls with centenarians. To calculate the observed effect sizes, we used logistic regression models correcting for population stratification (principal components 1-5). [18, 19] We calculated effect sizes and odds ratios relative to the least frequent allele assuming additive genetic effects, and estimated 95% confidence intervals. The change in effect size is then, for each SNP, the ratio between the observed effect size in each comparison and the reference effect size. When the change equals 1, the observed and reference effect sizes are the same; when the change is larger than 1, the observed effect size is larger than the reference effect; when the change is smaller than 1, yet positive, the reference effect size is larger than the observed effect; when the change is negative, the directions of the observed and the reference effect sizes are opposite.

## Supplementary Figures

**Figure S1:**
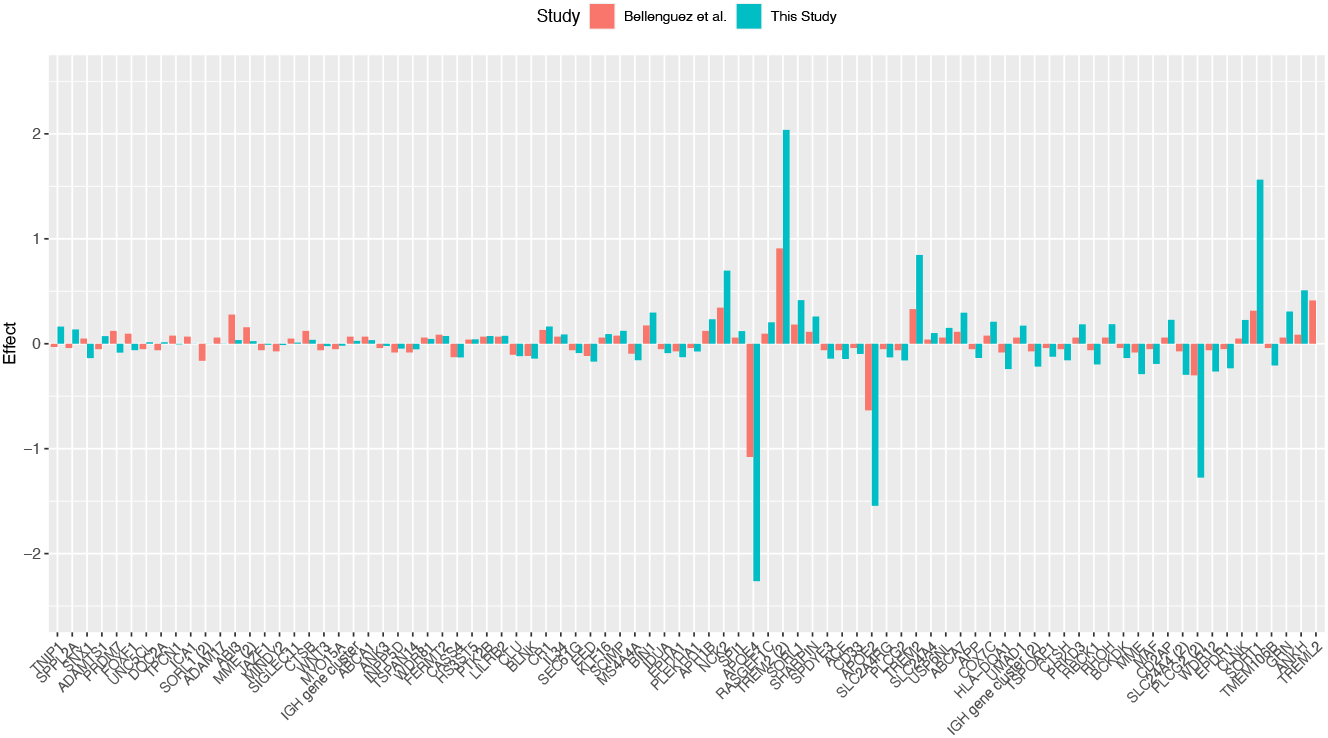
Single variant associations summary comparing AD cases and cognitively healthy centenarians. The figure shows the SNP effect sizes relative to the comparison of AD cases and cognitively healthy centenarians (blue bars) and as reported in the reference GWAS (red bars).

**Figure S2:**
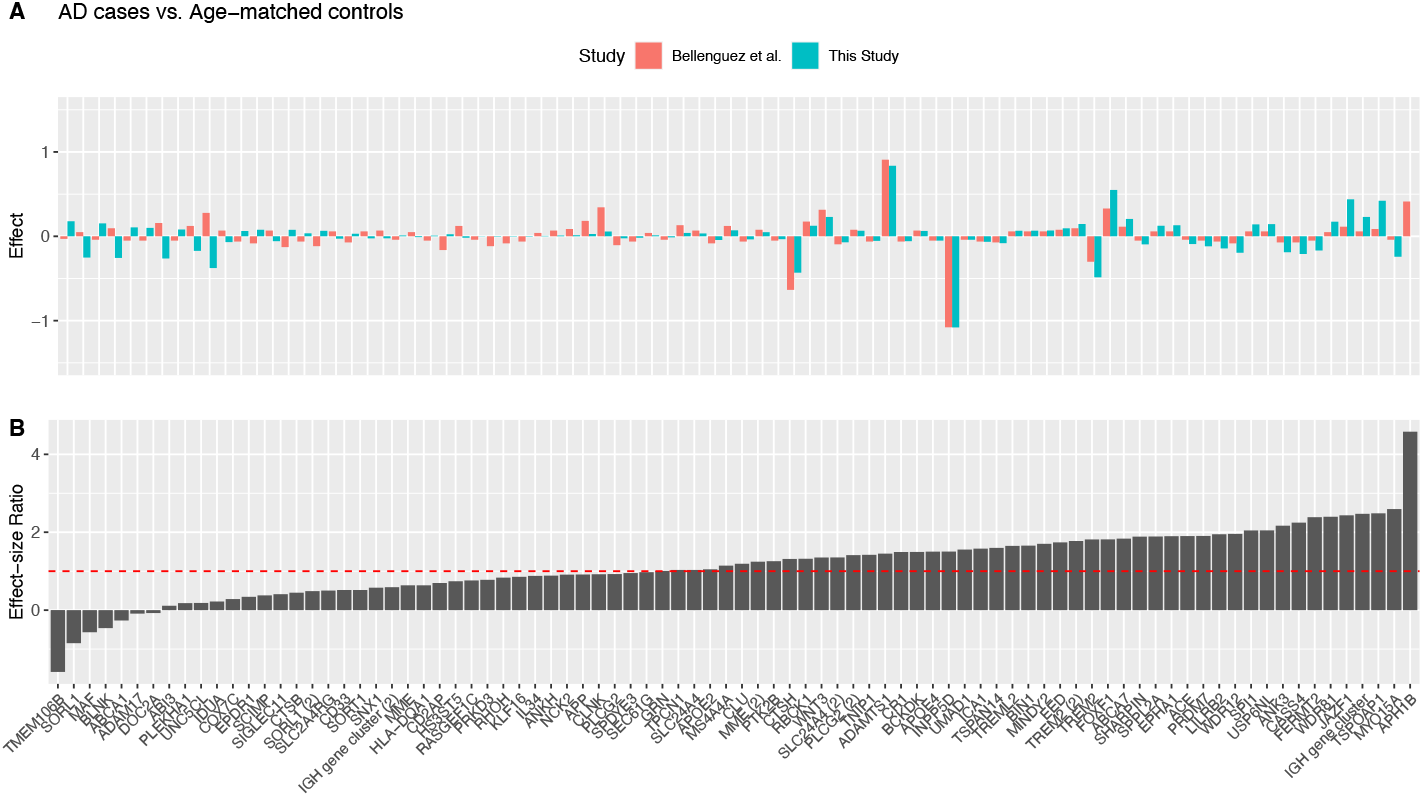
Single variant associations summary comparing AD cases and age-matched controls. Figure A shows the SNP effect sizes relative to the comparison of AD cases and normal controls (blue bars) and as reported in the reference GWAS (red bars). Figure B shows the change in effect size when comparing observed effect sizes (AD cases vs. normal age-matched controls) to the reference effect sizes. The dashed red line at 1 indicates the published effect size from the literature. Negative bars refer to a different direction of effect between the GWAS we used as a reference and our Study. Bars lower than 1 (yet positive), refer to SNP whole effect-size from the GWAS we used as a reference was larger than the observed effect-size.

**Figure S3:**
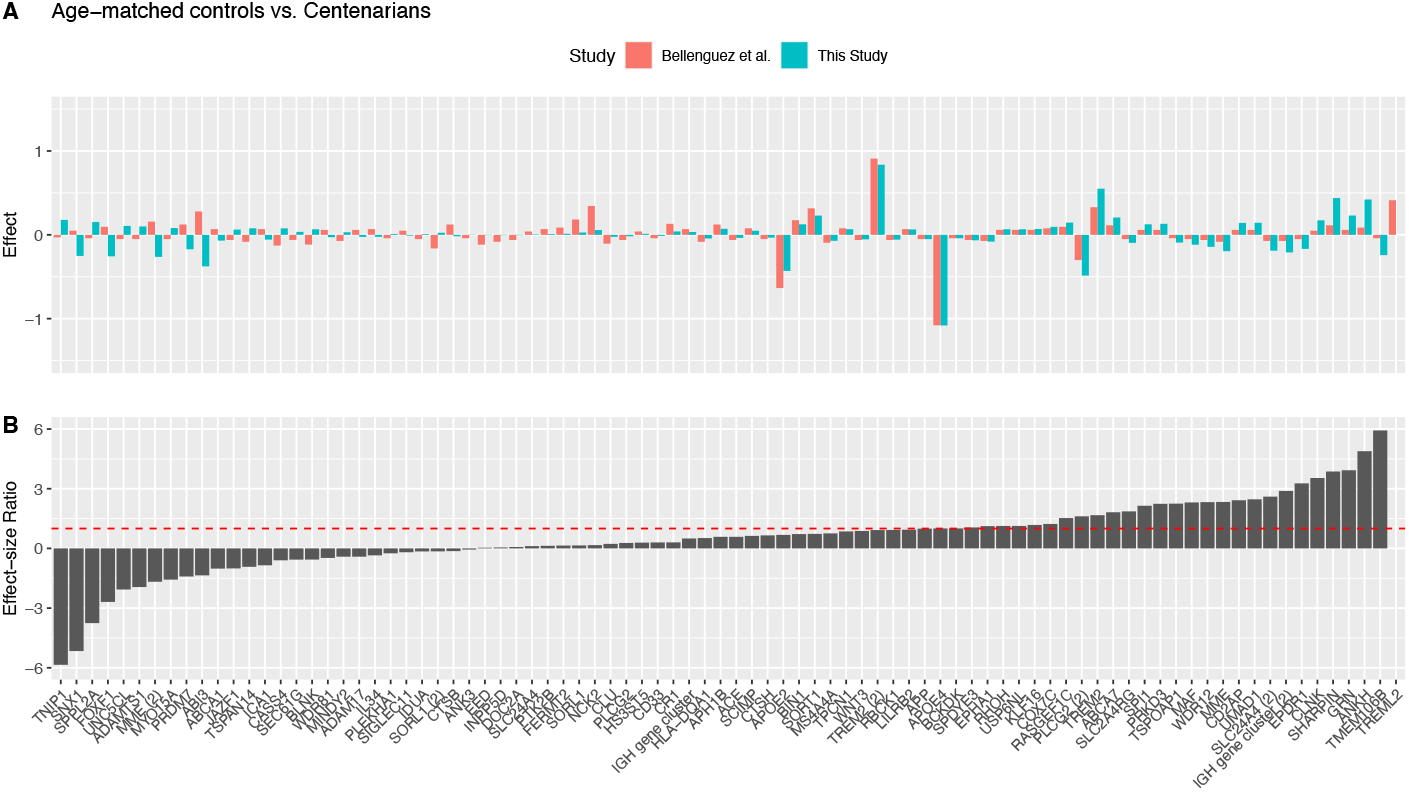
Single variant associations summary comparing healthy controls and cognitively healthy centenarians. Figure A shows the SNP effect sizes relative to the comparison of normal controls and centenarians (blue bars) and as reported in the reference GWAS (red bars). Figure B shows the change in effect size when comparing observed effect sizes (Normal controls vs. centenarians) to the reference effect sizes. The dashed red line at 1 indicates the published effect size from the literature. Only the association of the 2 SNPs in *APOE* remained significant after correcting for multiple tests. Negative bars refer to a different direction of effect between the GWAS we used as a reference and our Study. Bars lower than 1 (yet positive), refer to SNP whole effect-size from the GWAS we used as a reference was larger than the observed effect-size.

